# The COVID-19 Forecast in Northwest Syria The Imperative of Global Action to Avoid Catastrophe

**DOI:** 10.1101/2020.05.07.20085365

**Authors:** Mahmoud Hariri, Wael Obaid, Hazem Rihawi, Salah Safadi, Mary Ana McGlasson

## Abstract

**Introduction:** There is limited research on how the COVD-19 pandemic will affect countries with weakened health systems and particularly those in conflict. Syria’s protracted conflict has strained its health systems and caused fragmentation. In this study, we focus on northwest (NW) Syria, where recent violence has driven almost one million civilians (of the 4.17 million in the area) from their homes between December 2019 and March 2020. The area is challenged by overcrowding, inadequate WASH, shelter and insufficient healthcare services. Internationally promoted measures (social distancing, self-isolation, quarantine, lockdown) are not impossible. We model outcomes, according to three scenarios, should there be a COVD-19 outbreak. We aim to 1. Predict the numbers of cases, including severe and critical ones, and deaths. 2. Identify critical time points when the health system capacity is overwhelmed due to COVID-19.

**Methodology:** using the WHO COVD-19 Essential Supplies Forecasting Tool (COVID-ESFT) and data from the Health Information System Unit on population and health facility capacity and utilization in northwest Syria, we generate predicted numbers of cases, deaths and health care needs according to three scenarios. **Scenario One** assumes a medium doubling rate (every 4 days) and a medium clinical attack rate (20% of the population). **Scenario Two** assumes a *fast* doubling rate (every 3.2 days) and a medium clinical attack rate (20% of the population). **Camp-population Scenario** assumes a *very fast* doubling rate (every 2.3 days) and a medium clinical attack rate (20% of the population). Scenarios One and Two apply to the total population of 4.17 million and for 8 weeks from the first case while Camp-population Scenario applies only to the 1.2 million internally displaced persons (IDPs) in camps and tented settlements and for 6 weeks from the first case. For each scenario, we identify critical time-points when the health system capacity is overwhelmed assuming a highly conservative estimate that 50% of regular hospital (ward) and ICU beds can be occupied by COVID-19 patients.

**Results:** **Scenario One** predicts 16,384 cases (0.4% of the total population), of which 2,458 are severe and 819 are critical, and 978 deaths in the first 8 weeks. **Scenario Two** predicts 185,364 cases (4.4% of the population), of which 27805 are severe and 9268 are critical, and 11,066 deaths in the first 8 weeks. **Camp-population Scenario** predicts 240,000 cases (20% of the IDP population) of which 36,000 are severe and 12,000 are critical and 14,328 deaths in the first 6 weeks. With only 2,429 inpatient beds and 240 ICU beds (98 with adult ventilators, 62 with paediatric ventilators) in northwest Syria, ward and ICU bed capacities will be overwhelmed within 4-7 weeks. The Camp-population Scenario will see the earliest critical time-points.

**Conclusion and recommendations:** Should a COVID-19 outbreak occur in NW Syria, projected cases and deaths will be particularly severe among IDPs. Health system capacity will be overwhelmed within a short period after the first case in camp settings. There is need for further research to account for additional variables that can impact projections. However, it is urgent for international community to mobilize efforts and resources to support community-based measures, increase testing, strengthen health system capacity.

## Introduction

Cases of an unusual pneumonia in Wuhan city in Hubai Province of China first came to the attention of WHO at the end of December 2019. Since then, there have been 3.04 million confirmed cases and 211,000 deaths (as of 28^th^ April 2020.).^1^

The response to the COVID-19 pandemic has proven challenging even for some high-income countries whose health systems are more robust than those in low- and middle-income countries, particularly those affected by protracted conflicts or humanitarian crises. Most published research has focused on high income countries with many unknowns remaining for how a COVD-19 outbreak will affect those countries with less robust health systems. Concerns are that countries like Syria will face challenges to the healthcare system which may be rapidly overwhelmed.

Syria’s protracted conflict has strained its health systems across the country; in fact, there is more than one health system functioning within the country e.g. areas under government control, NE Syria, NW Syria and areas under Turkish control. In this report, we will focus on NW Syria where a further escalation of violence has driving almost one million civilians (of the 4.17 million in the areas) from their homes between December 2019 and March 2020. The area is challenged by overcrowding, inadequate WASH, inadequate shelter and insufficient healthcare services. In these areas, internationally promoted measures (social distancing, self-isolation, quarantine, lockdown) are nigh impossible.

The proportion of severe cases (e.g. dyspnea, hypoxia, >50% lung involvement on imaging – requiring hospital admission) and critical cases (respiratory failure, shock, multi-organ failure – requiring ICU admission) and mortality differ in different contexts. As such, it remains unclear on what proportions will occur among the population in NW Syria in either the population in the internally displaced people (IDP) camps where 1.2 million of the 4.17 million population in NW Syria reside or among other residents in the area. Age and comorbidities (e.g. hypertension, diabetes) can result in more severe disease. In this area, it is estimated that there 76,000 people aged 60 years or over and up to 40% of the adult population have a comorbidity.

Mortality estimates vary by setting and are likely affected by the effectiveness and availability of testing. The Case Fatality Rate (CFR) in Wuhan was 5.8% and in Italy it was 7.2% in mid-March 2020;^2^ Italy’s current CFR is above 13%^3^. Global mortality is between 6-7%, and COVID-19 cases have continued to rise across the world. Mortality in areas of protracted conflict like Syria are as yet unknown but may be higher despite the lower median age of the population due to other factors e.g. poor nutrition, illness, poor healthcare system access.

As of 27^th^ April 2020, Syria reports 42 confirmed cases (3 deaths) in GCAs and there is one confirmed case of a person who died in NE Syria. Of 216 tests performed in NW Syria (as of 19^th^ April 2020), all have been negative. There is concern about widespread under-testing and under-reporting given Syria’s proximity to countries which among the top ten for COVD-19 cases internationally, particularly Turkey and Iran who have 112,000 and 92,584 cases respectively (28^th^ April 2020.)

## Population and risk factors

The Humanitarian Needs Assessment Program estimates that the population in NW Syria is 4,170,000 of whom 67% are IDPs; 1.2 million are living in tented settlements or camps. The population in NW Syria is particularly vulnerable to COVID-19 spread given ongoing hostilities, wide-spread displacement, limited WASH services and inadequate shelter. Almost 1 million civilians were displaced by the escalation of hostilities between December 2019 and March 2020.

Though the population in NW Syria is skewed towards a younger demographic with fewer patients aged 60 years or over (see population pyramid) the population has a number of other risk factors. Up to 41% of the adult Syrian population has a non-communicable disease e.g. hypertension, diabetes, cancer;^4^. Smoking prevalence among Syrian adults (particularly men) is among the highest in the Middle East and is associated with more severe disease.^5^ Malnutrition (micro and macronutrient deficiency) is high given food insecurity in the area.^6^ These may affect the proportion who develop severe or critical disease and could potentially contribute to a more rapid spread of COVID-19 than seen elsewhere with a higher doubling rate, higher clinical attack rate and a high proportion of patients requiring hospitalization or ventilation than in other settings.

## Health System Status

Syria has suffered more than 9 years of conflict and its health system has been affected by both direct and indirect attacks on healthcare. The conflict has affected every aspect of the healthcare system. As of April 2020, 306 health facilities remain functional out of 568 health facilities in NW Syria^8^ which is insufficient for the needs of the population. Healthcare worker shortages have resulted from the forced displacement of healthcare workers (around 67% of Syria’s total healthcare workforce has been displaced outside of Syria) and more than 932 healthcare workers have been killed during the conflict^9^ (see figure 2) This has contributed to significant healthcare system and healthcare worker gaps which are unable to meet the needs of the population, even prior to the threat of COVID-19. (see figure 3)

**Figure 1.**
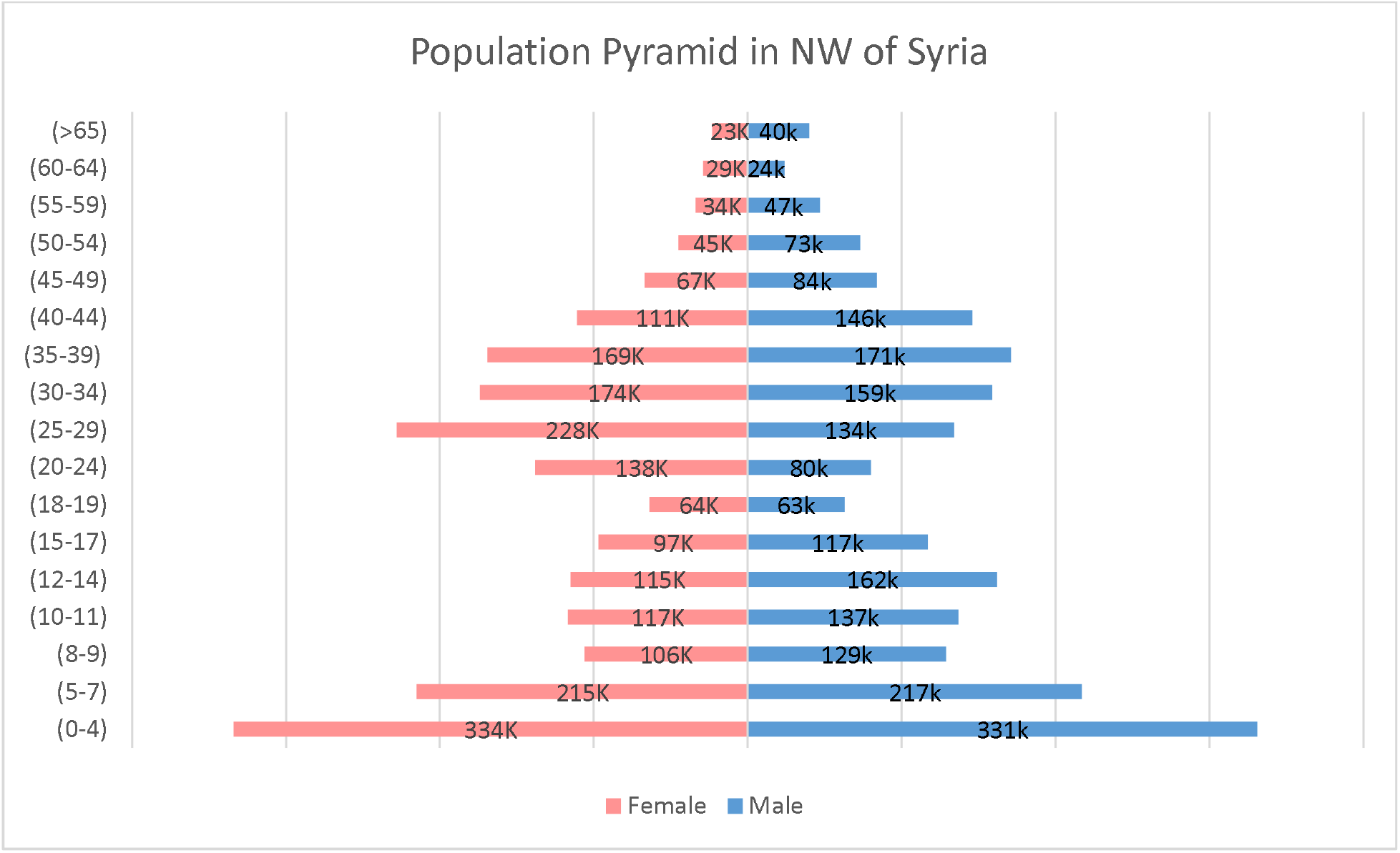
Population distribution in NW Syria^7^

**Figure 2.**
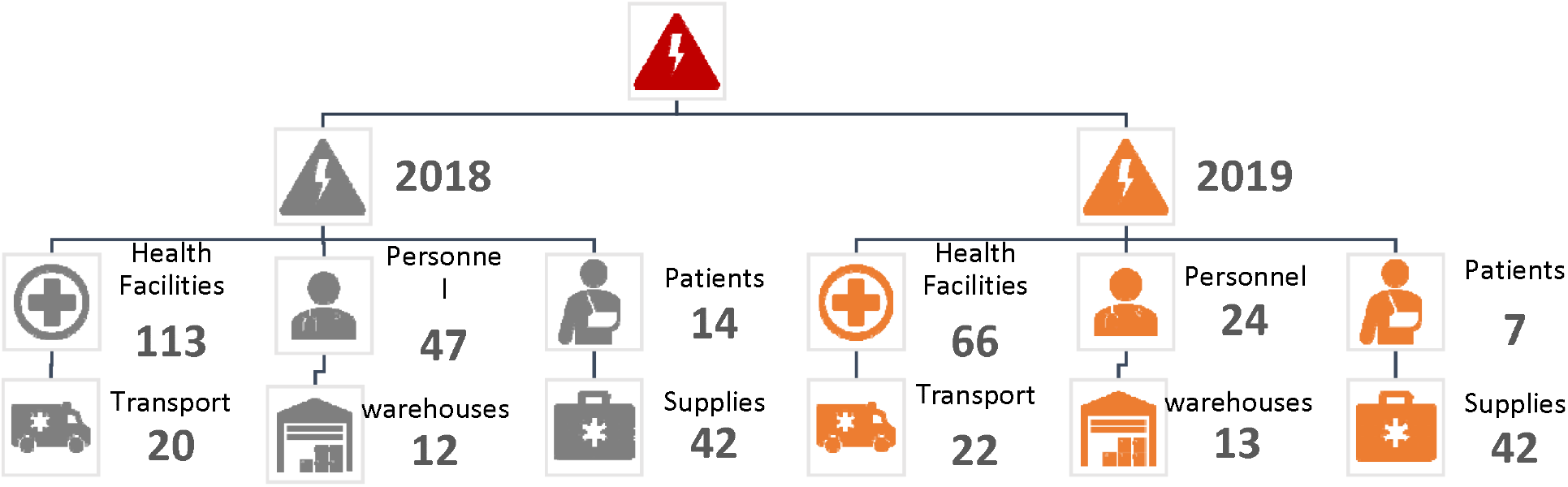
Attacks on health care facilities across Syria in 2018 (Black) and 2019 (orange)^14^

**Figure 3.**
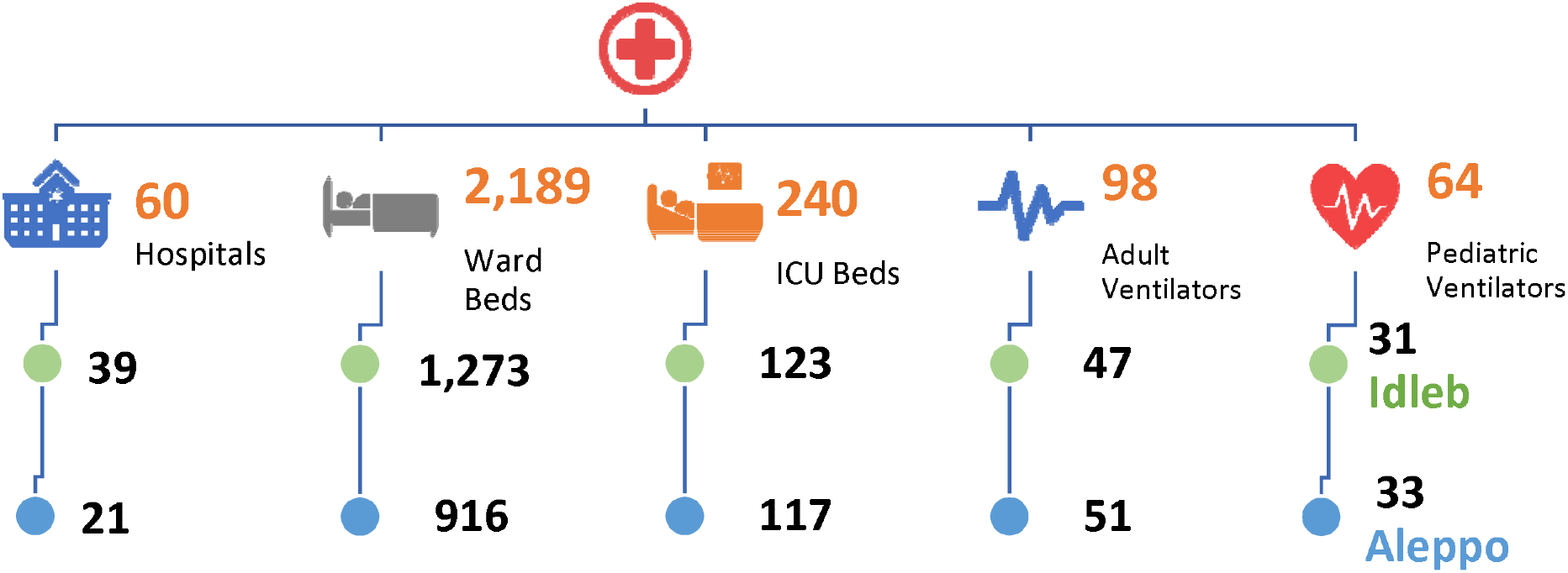
Available hospitals and their capacities in NW Syria as of April 2020^15^

According to the local Health Information System Unit^10^ there are 2,429 beds including 240 ICU beds (98 with adult ventilators and 62 with paediatric ventilators). The mean 2019 ward occupancy percentage was 88% and ICU occupation was 98%^11^. The Global Health Cluster suggests in its core indicators and benchmarks that there should be at least 10 inpatient hospital beds per 10,000 people (inpatients & maternity) and that 10% of these should be ICU beds. This means that for the NW Syria population of 4,170,000, there should ideally be more than 4,170 beds and 417 ICU beds to meet minimal needs of the NW Syrian population for normal operation. As such, even prior to the COVID-19 pandemic there was almost a 50% shortfall for inpatient beds.

In terms of available healthcare workers, the highest estimates locally available suggest there are about 1,000 doctors and 2,000 nurses and midwives remaining in NW Syria, which equals to 12 eight healthcare workers per 10,000 people^12^. This falls far below the SPHERE indicator of >22 per 10,000 population^13^. Beyond this, there are insufficient numbers of healthcare workers with the right skills to manage ICU patients and operate ventilators for the expected rise in COVID-19 critical cases. Furthermore, in NW Syria there is no unified governing body following years of ongoing conflict. The main health governance body is the Idleb Health Directorate, while in northern Aleppo, there are multiple local health authorities including the Aleppo Health Directorate and the Syrian-Turkish Task Force.

## Aim & Objectives

The overall aim of this study is to assist decision makers to prepare the local healthcare system (community isolation beds, inpatient facilities, ICU beds, human resources, testing and triage facilities) to meet the needs of the population in the likely scenario that cases of COVID-19 are detected in NW Syria. The objectives are to: Generate estimates of projected case load, by severity, deaths and health system needs over the first 8 weeks of an epidemic according to various scenarios affecting the whole population or the highly-vulnerable population of camp dwellers; identify critical time-points at which the health system capacity to manage COVID-19 cases is exceeded and the health system is at risk of collapse; and use these findings to draw recommendations to support ongoing preparedness planning

## Methodology

This study uses the WHO COVID-ESFT and data from the Health Information System Unit on population, health system capacity and utilization for NW Syria to build a model of potential numbers of cases and deaths due to COVID-19 and to anticipate critical time-points when the health system would be overwhelmed as number of severe and critical cases exceed number of available ward and ICU beds, respectively. We generate predicted numbers of cases, deaths and health care needs according to three scenarios. ***A****ll-population Scenario One* and *Two* apply to the total population of 4.17 million and for 8 weeks from the first case. They differ in that Scenario One assumes a medium doubling rate (every 4 days), and is thus more optimistic, while Scenario Two assumes a *fast* doubling rate (every 3.2 days) which is less optimistic. *Camp-population Scenario* only to the 1.2 million internally displaced persons (IDPs) in camps and tented settlements along the Turkey-Syria border. This scenario predicts a bad-case scenario in a very crowded population with limited access to social distancing, appropriate shelter and adequate WASH. The scenario assumes a *very fast* doubling rate (every 2.3 days, the highest available rate in the tool). The COVID-ESFT has a limitation with such parameters, whereby it can only forecast a six-week timeframe. The key parameters of each model are captured in Table 1.

**Table 1.** The key parameters

| Parameter | All-population Scenario One | All-population Scenario Two | Camp-population Scenario |
| --- | --- | --- | --- |
| Known Cumulative Cases (#) | 1 | 1 | 1 |
| Estimation Method | Exponential Growth | Exponential Growth | Exponential Growth |
| Doubling Rate (days) | Medium (4 days) | Fast (every 3.2 days) | Very fast (every 2.3 days) |
| Clinical Attack Rate | Medium (20% of population) | Medium (20% of population) | Medium (20% of population) |
| Testing Strategy | Targeted | Targeted | Targeted |

**Table 2.** Scenario One prediction

| Week | Cases | Mild Cases | Severe Cases | Critical Cases | Deaths |
| --- | --- | --- | --- | --- | --- |
| Week 1 | 3 | 3 | 1 | - | - |
| Week 2 | 11 | 9 | 2 | 1 | 1 |
| Week 3 | 38 | 30 | 6 | 2 | 2 |
| Week 4 | 128 | 102 | 19 | 6 | 8 |
| Week 5 | 431 | 344 | 65 | 22 | 26 |
| Week 6 | 1,448 | 1,159 | 217 | 72 | 86 |
| Week 7 | 4,871 | 3,897 | 731 | 244 | 291 |
| Week 8 | 16,384 | 13,107 | 2,458 | 819 | 978 |

➢ **Cumulative Known Cases:** NW Syria has not yet confirmed the first COVID-19 case. The COVID-ESFT advises to enter number one as minimum, which corresponds with <u>***week*** 1</u> of the outbreak, based on this equation:

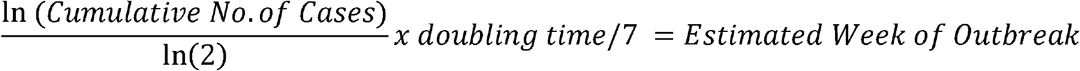
➢ **Cumulative Case Estimation Method:** The COVID-ESFT suggests using “exponential growth” in most cases, which is what was selected for each of the three scenarios. The exponential growth equation to calculate the cumulative number of COVID-19 cases for each week until the estimated end of the epidemic is as follows:

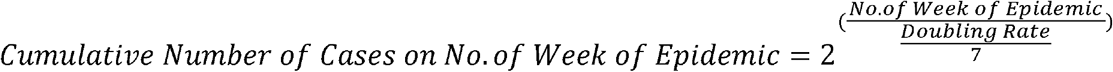
➢ **Doubling Rate (Days):** This element of the tool estimates the number of days at which the cases are expected to double. This study used three doubling rates as follows^16^,^17^:
  - **Scenario One**: *medium* doubling rate (every 4 days) (Population: NW Syria population of 4.17 million)
  - **Scenario Two**: *fast* doubling rate (every 3.2 days) (Population: NW Syria population of 4.17 million)
  - **Camp-population Scenario**: *very fast* doubling rate (every 2.3 days) due to the very dense and vulnerable population of IDPs within the camps (Population: 1.2 million people who are in camps)
➢ **Clinical Attack Rate:** “Attack rate is defined as the proportion of the population at risk that gets symptomatic after exposure to the novel Coronavirus-19 during the period of the epidemic.”. This study used a medium clinical attack rate for all scenarios, which means 20% of the population will be infected and become symptomatic. We assumed this estimate for purposes of modeling based “that the clinical attack rate was 20% in the first year of the 2009 H1N1 pandemic”^18^, but the reality could be worse. Some studies suggest clinical attack rates could reach as high as 50-80%^19^.
➢ **Targeted Testing Strategy:** In all scenarios, we modelled based on a targeted testing strategy. This means that severe and critical cases are prioritized over other patients, which meet the WHO case definition.
➢ **Timeframe**: For each scenario, we observed COVID-19 estimated data in terms of cumulative cases, surveillance and lab needs, severe cases, critical cases and expected case fatality rate for 8 weeks from the first diagnosed case.
➢ **Case fatality rate**: We use the global average CFR, estimated at 5.9 % based on available data on 11 April^20^, to project deaths.
➢ **Severe and critical cases**: Prior reports, including from the Chinese Centers for Disease Control and Prevention, suggest that 14% of COVID-19 cases will be severe and 5% will be critical^21^ Said differently, about 33% of the severe cases are projected to be critical. We use these estimates, in all the three scenarios, to generate numbers of predicted severe and critical cases.
➢ **Defining critical time-point in health system capacity:** The researchers considered that the health system capacity reaches critical time-points when number of severe and critical COVID-19 patients requiring hospitalization reaches 50% of number of all available ward and ICU beds, respectively. Considering the total number of ward beds (2,148) and ICU beds (240), this translates to 1,100 ward beds and 120 ICU beds.

## Results

### All-population Scenario One

<u>Total Cases:</u> Within the first eight weeks, the COVID-ESFT projects a total of 16,384 cases, equating to 0.4% of the total population of 4,170,000, and 978 deaths. These estimates can be broken down by week and case severity as depicted in Table 3 and Figure 4. Scenario One shows a significant increase in the total number of the cases starting from week 6.

**Table 3.** Scenario Two prediction

| Week | Cases | Mild Cases | Severe Cases | Critical Cases | Deaths |
| --- | --- | --- | --- | --- | --- |
| Week 1 | 5 | 4 | 1 | - | - |
| Week 2 | 21 | 17 | 3 | 1 | 1 |
| Week 3 | 95 | 76 | 14 | 5 | 6 |
| Week 4 | 431 | 344 | 65 | 22 | 26 |
| Week 5 | 1,961 | 1,569 | 294 | 98 | 117 |
| Week 6 | 8,933 | 7,147 | 1,340 | 447 | 533 |
| Week 7 | 40,693 | 32,555 | 6,104 | 2,035 | 2,429 |
| Week 8 | 185,364 | 148,291 | 27,805 | 9,268 | 11,066 |

**Figure 4.**
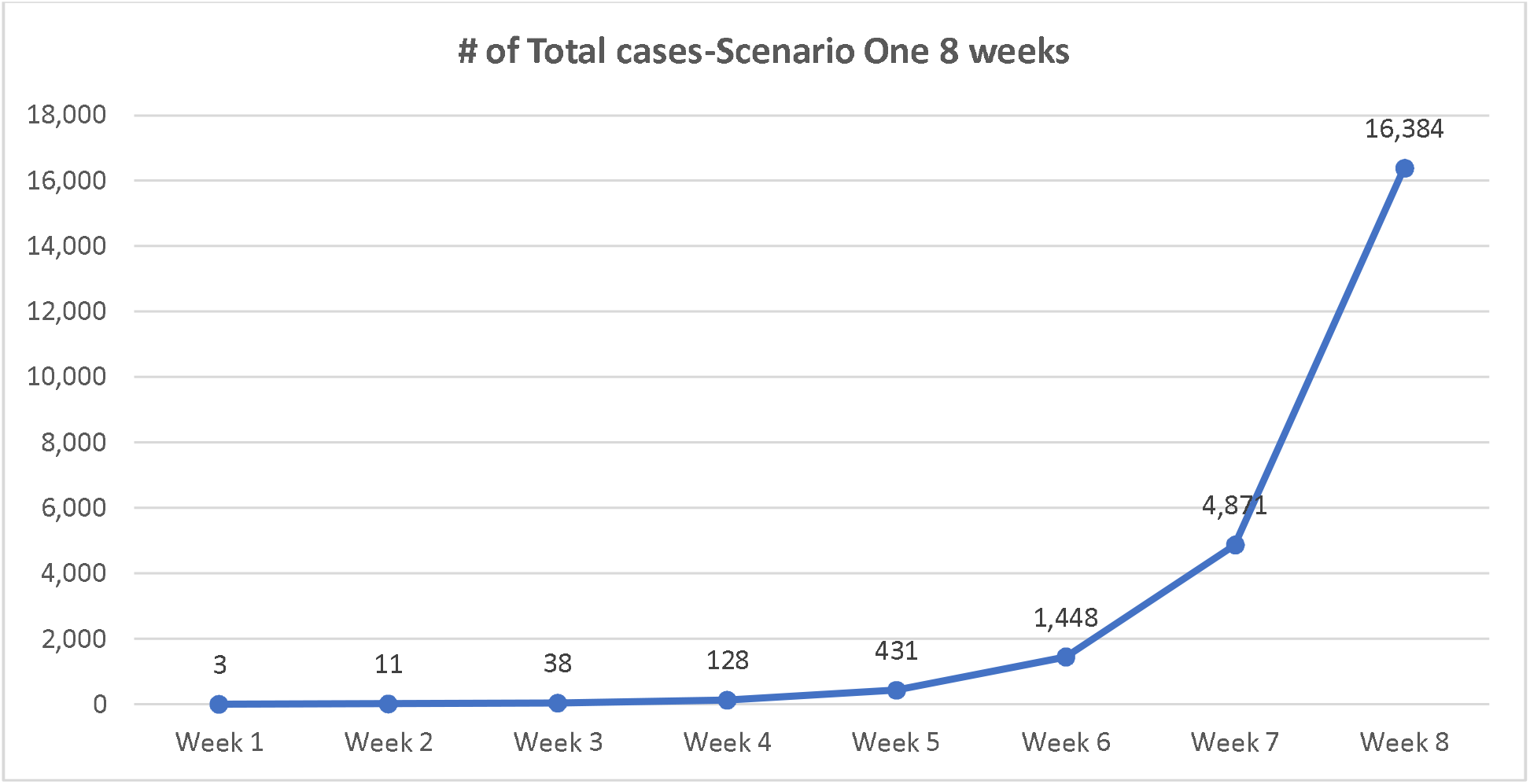
Scenario One predicted total cases

**Figure 5.**
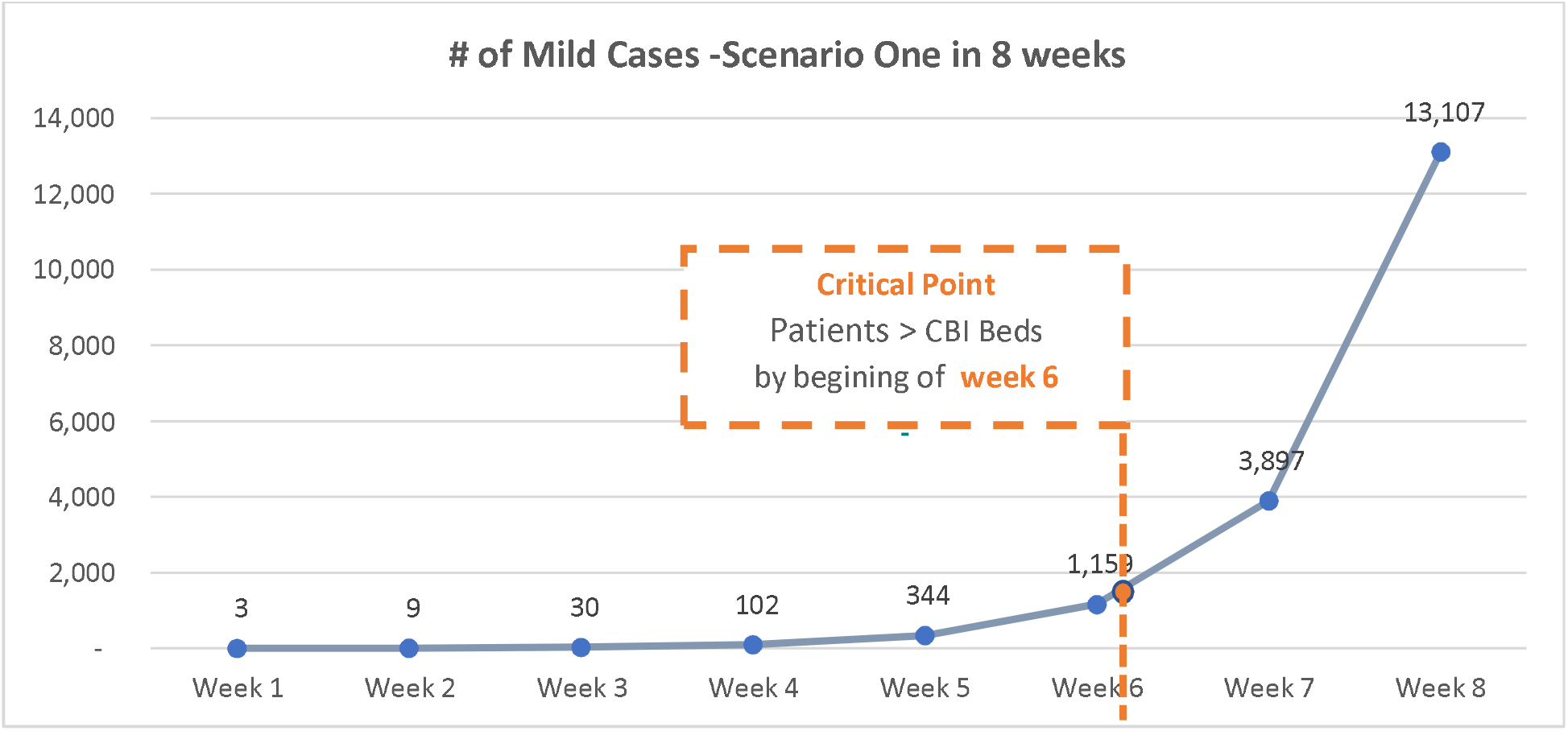
Scenario One predicted mild cases

**Figure 6.**
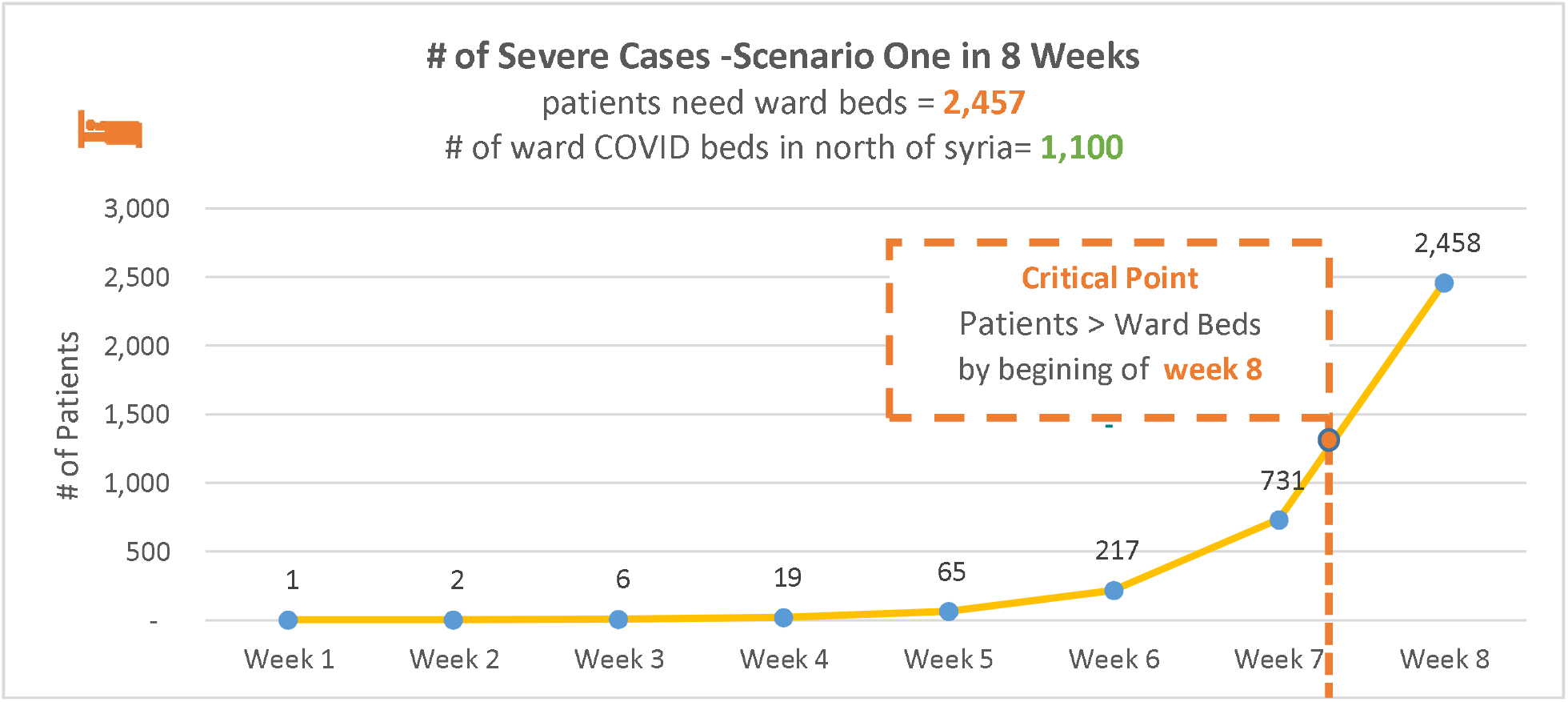
Scenario One predicted severe cases

**Figure 7.**
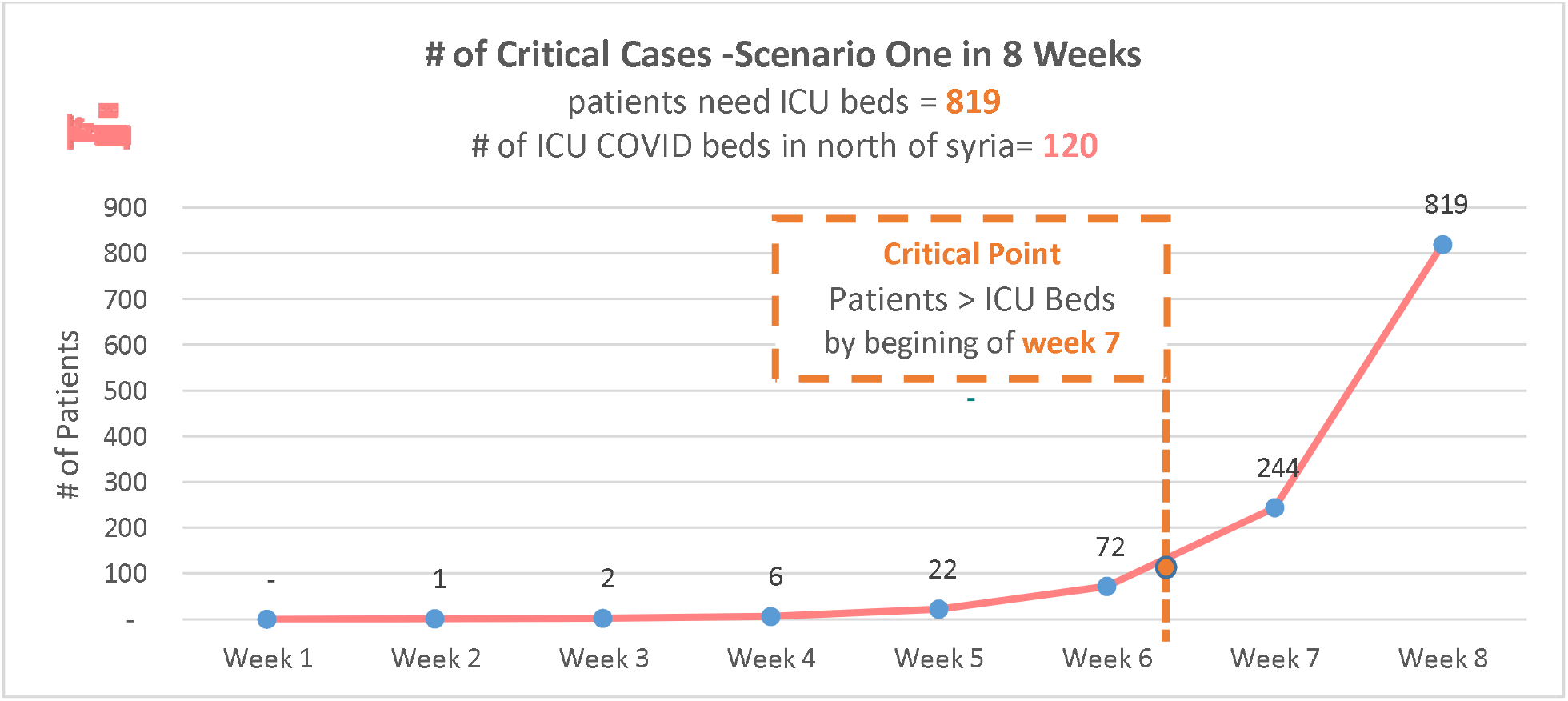
Scenario One predicted critical cases

**Figure 8.**
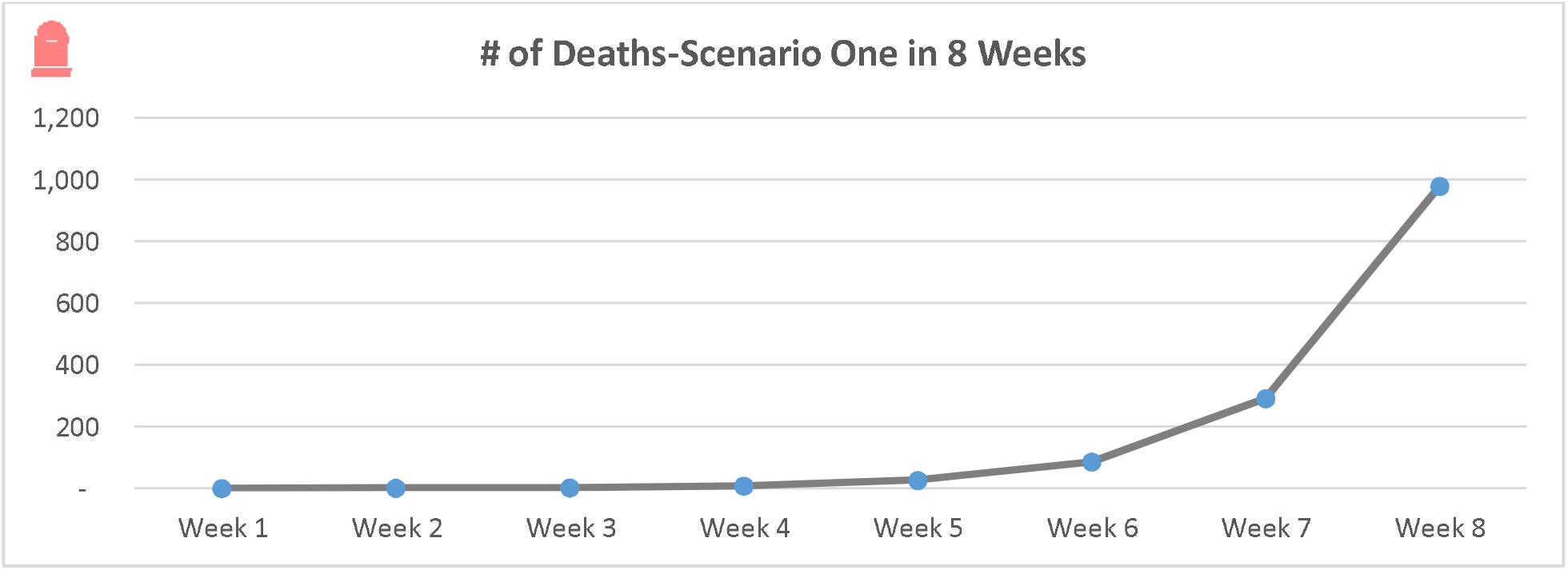
Scenario One predicted deaths

<u>Mild Cases:</u> By the end of the 8 -week forecasting period, COVID-ESFT predicts 13,107 mild cases that need to be identified and isolated. These cases could be managed in community-based isolation (CBI). CBI is necessary due to the high-density and poor living conditions of the population where self-isolation and social distancing would not be possible. Currently there are plans to establish 1,400 beds distributed over 30 CBI areas. These beds will be saturated after 6 weeks taking into consideration the need for a two-week isolation period per patient.

<u>Severe Cases:</u> Out of total 16,384 predicted cases, 2,458 patients (14%) are expected to be severe, requiring inpatient ward admission. Needs for ward beds surpass available ward beds at the beginning of week 8, overwhelming the health system and potentially leading to increases in the number of critical cases which could contribute to a higher mortality rate.

*The health system in NW Syria would be unable to cope by the beginning of week 8; as such, severe cases could become critical and mortality could increase*

<u>Critical Cases:</u> Over 8 weeks, 819 cases (5% of all cases) will be critical, needing ICU admission. Needs for ICU beds surpass available ward beds at the beginning of week 7, overwhelming the health system and potentially leading to higher mortality.

*The critical point for ICU admissions capacity is expected during the seventh week*.

<u>Deaths:</u> The total number of expected COVID-19 deaths is 978. However, it could be much greater should the needs of the cases classified as severe or critical not be met in ward beds or ICU beds. This could increase the number of deaths.

### All-population Scenario Two

<u>Total Cases:</u> Within the first eight weeks, the COVID-ESFT projects a total of 185,364 cases, equating to 4.4% of the total population of 4,170,000, and 11,066 deaths. These estimates can be broken down by week and case severity (Table 4 and Figure 9). The model predicts a significant increase in cumulative cases starting after week 6.

**Table 4.** Scenario three prediction

| Week | Cases | Mild Cases | Severe Cases | Critical Cases | Deaths |
| --- | --- | --- | --- | --- | --- |
| Week 1 | 8 | 7 | 1 | - | - |
| Week 2 | 68 | 54 | 10 | 3 | 4 |
| Week 3 | 560 | 448 | 84 | 28 | 33 |
| Week 4 | 4,621 | 3,697 | 693 | 231 | 276 |
| Week 5 | 38,097 | 30,478 | 5,715 | 1,905 | 2,274 |
| Week 6 | 240,000 | 192,000 | 36,000 | 12,000 | 14,328 |

| **Week** | **Cases** | **Mild Cases** | **Severe Cases** | **Critical Cases** | **Deaths** |
| --- | --- | --- | --- | --- | --- |
| **Week 1** | 8 | 7 | 1 | - | - |
| **Week 2** | 68 | 54 | 10 | 3 | 4 |
| **Week 3** | 560 | 448 | 84 | 28 | 33 |
| **Week 4** | 4,621 | 3,697 | 693 | 231 | 276 |
| **Week 5** | 38,097 | 30,478 | 5,715 | 1,905 | 2,274 |
| **Week 6** | 240,000 | 192,000 | 36,000 | 12,000 | 14,328 |

**Figure 9.**
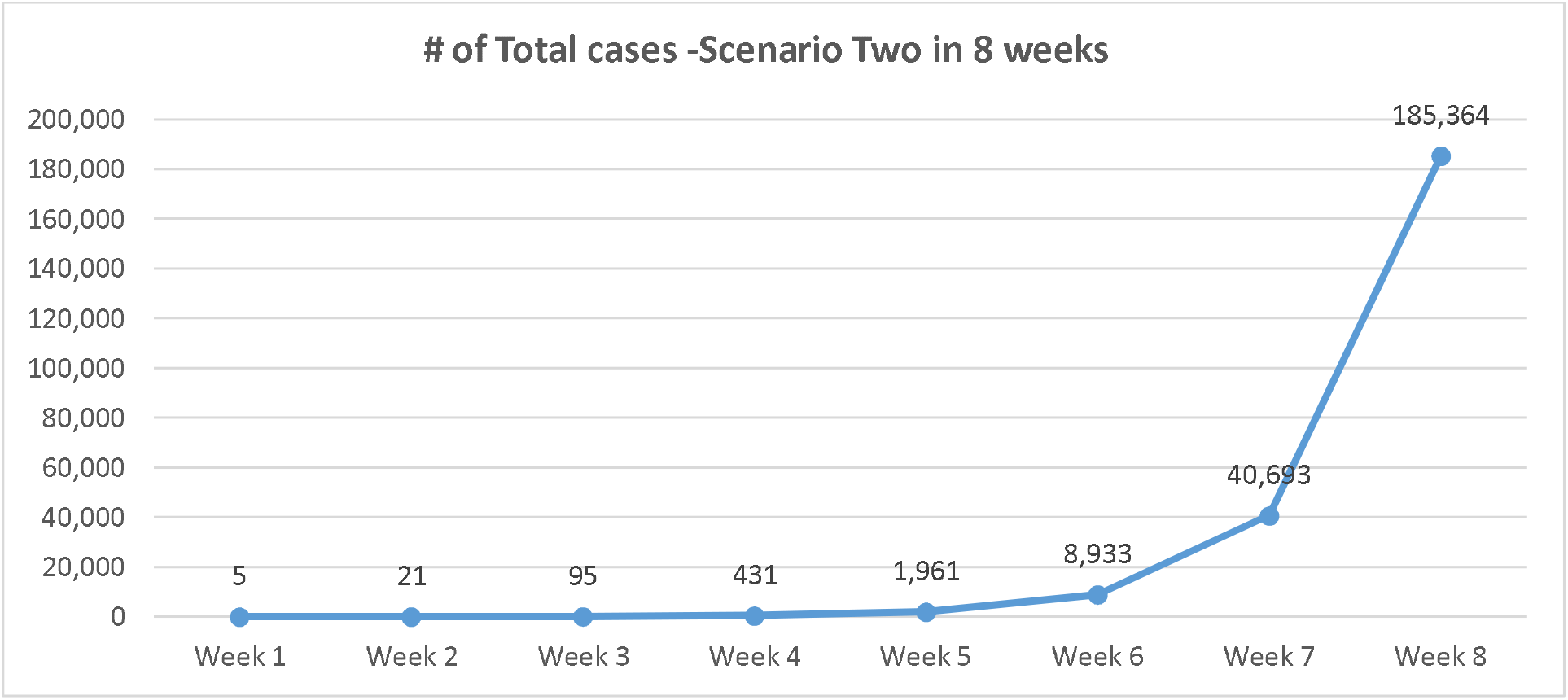
Scenario Two predicted Total cases

**Figure 10.**
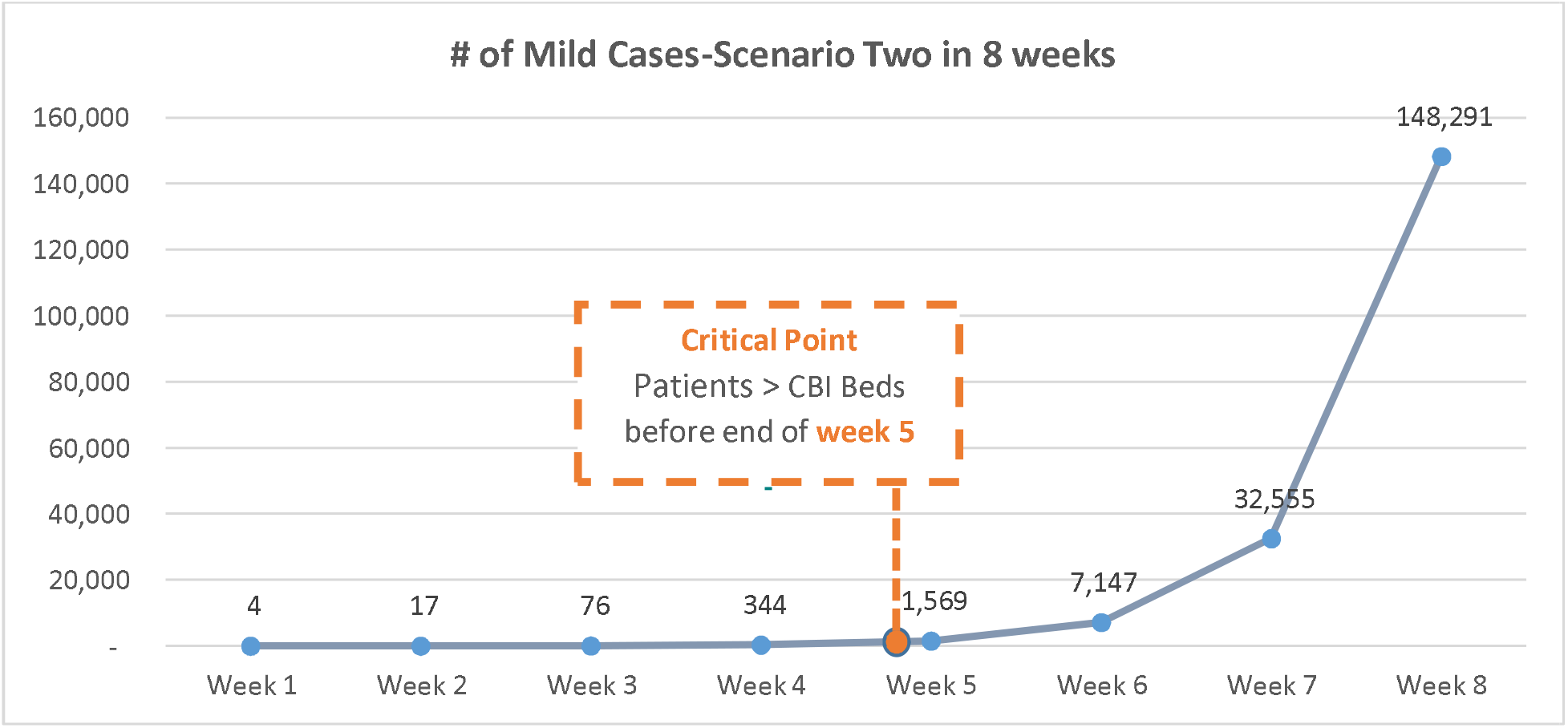
Scenario Two predicted mild cases

**Figure 11.**
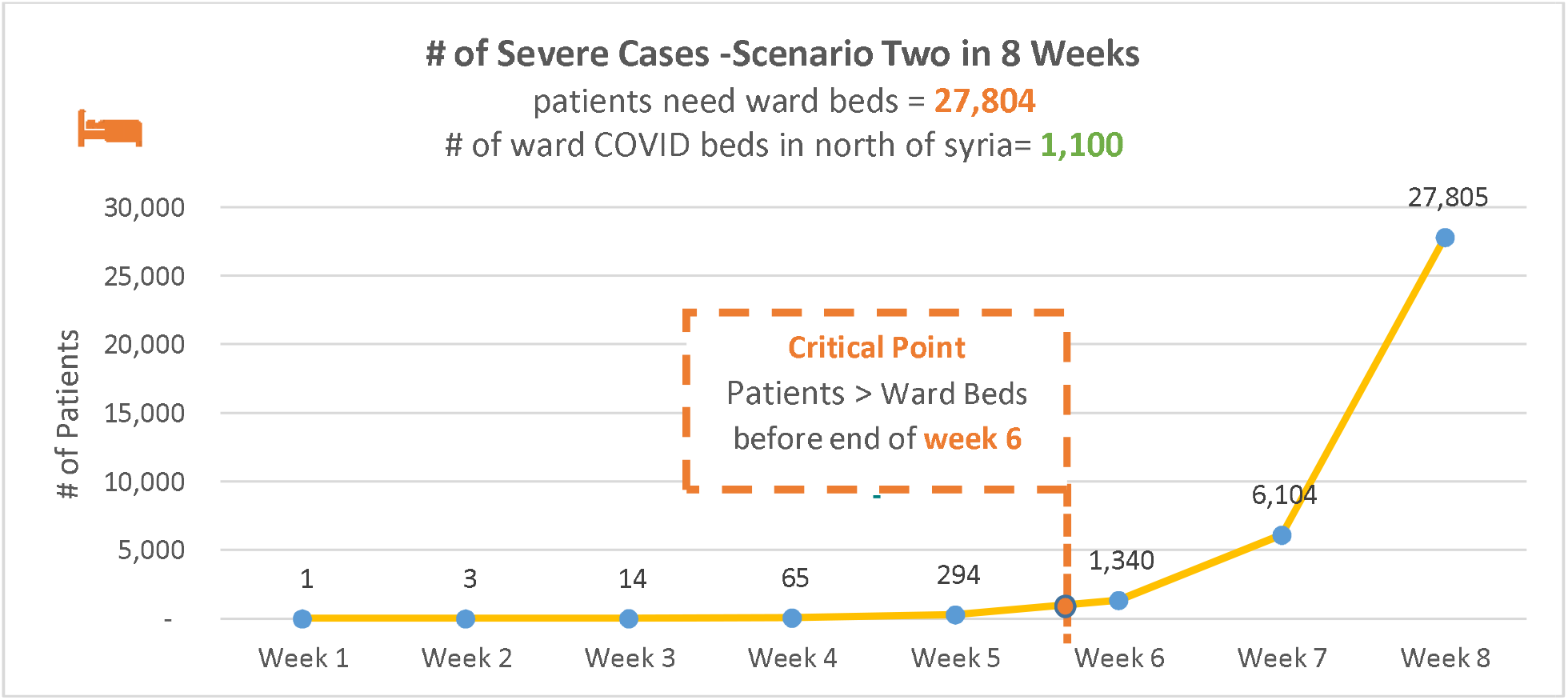
Scenario two predicted severe cases

**Figure 12.**
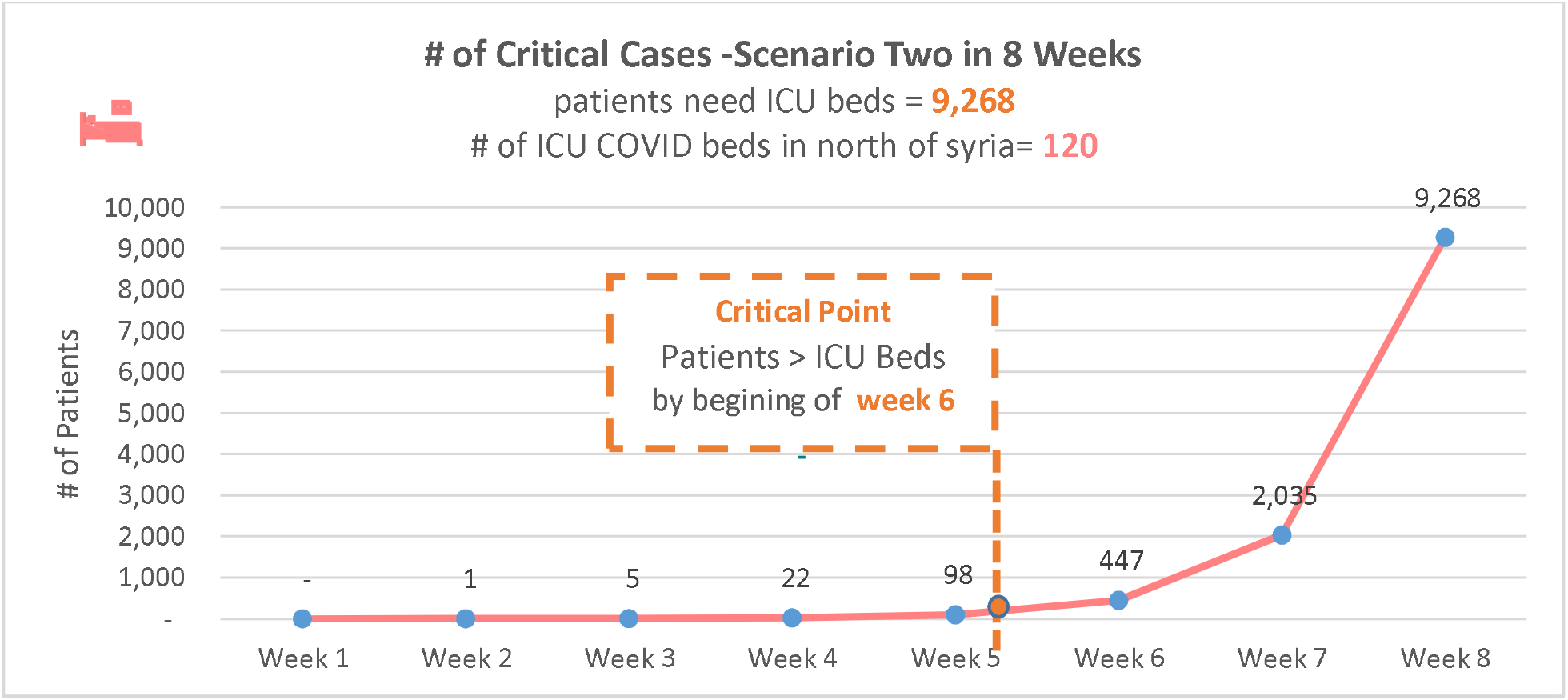
Scenario Two critical cases

**Figure 13.**
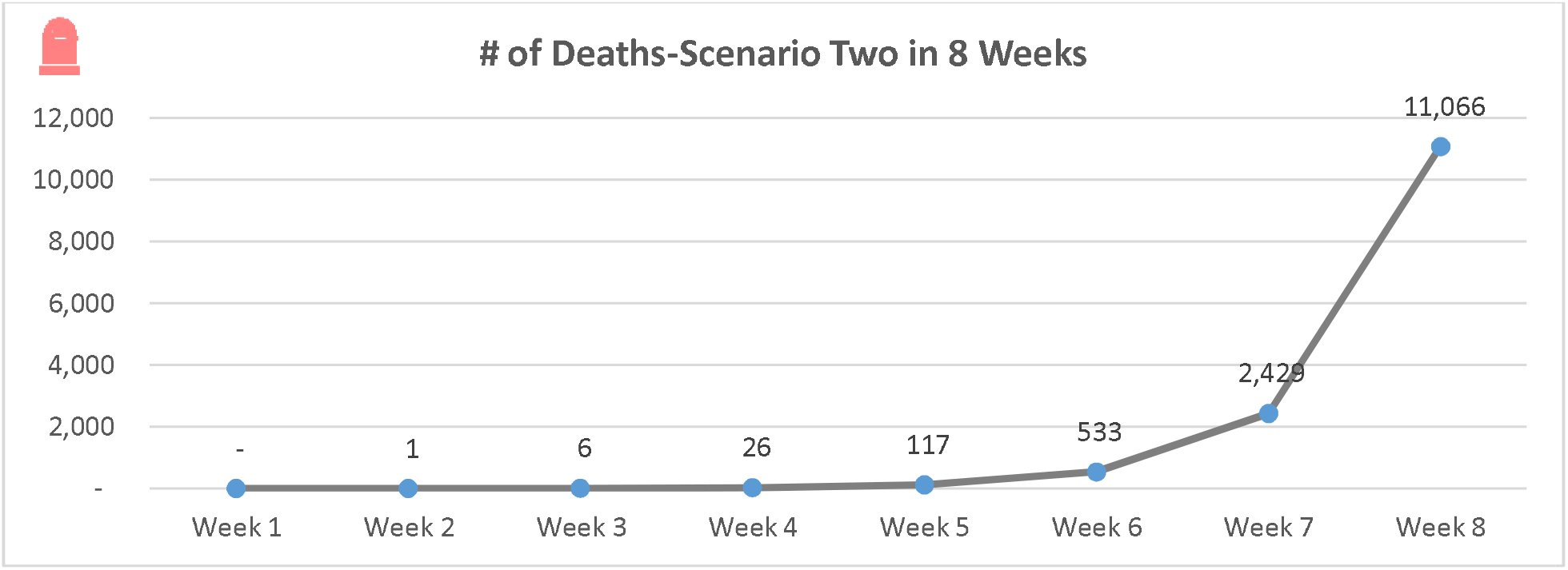
Scenario two predicted deaths

<u>Mild Cases:</u> By the end of the 6 -week forecasting period, COVID-ESFT predicts 148,291 mild cases that need to be identified and isolated. These cases could be also managed in CBI but CBI beds will be saturated before the end of fifth week considering the two weeks isolation period needed per patient.

<u>Severe Cases:</u> The COVID-ESFT predicts 27,804 severe cases out of the total 185,359 cases. These cases need inpatient admission at secondary health care centers but not ICU admission or ventilation. Needs for ward beds surpass available ward beds before the end of week 6, overwhelming the health system and potentially leading to increases in the number of critical cases which could contribute to a higher mortality rate.

<u>Critical Cases:</u> Over 8 weeks, 9,268 critical cases are estimated to need ICU support. The critical moment for critical cases occurs by the beginning of week 6, when ICU COVID-19 needs exceed 50% of the available ICU bed capacity.

<u>Deaths:</u> The COVID-ESFT predicts 11,066 deaths by the end of week 8. However, both the CFR and the number of deaths are expected to be much higher due to lack of health system capacity to manage the predicted number of severe and critical cases.

### Camp-population Scenario

<u>Total cases:</u> Within the first six weeks, the COVID-ESFT projects a total cumulative case load of 240,000, equating 20% of the total population of the camps (1,200,000), and 18,751 deaths. These estimates can be broken down by week and case severity (Table 6 and Figure 14). There will be a significant increase in cumulative number of cases starting from week 4.

**Figure 14.**
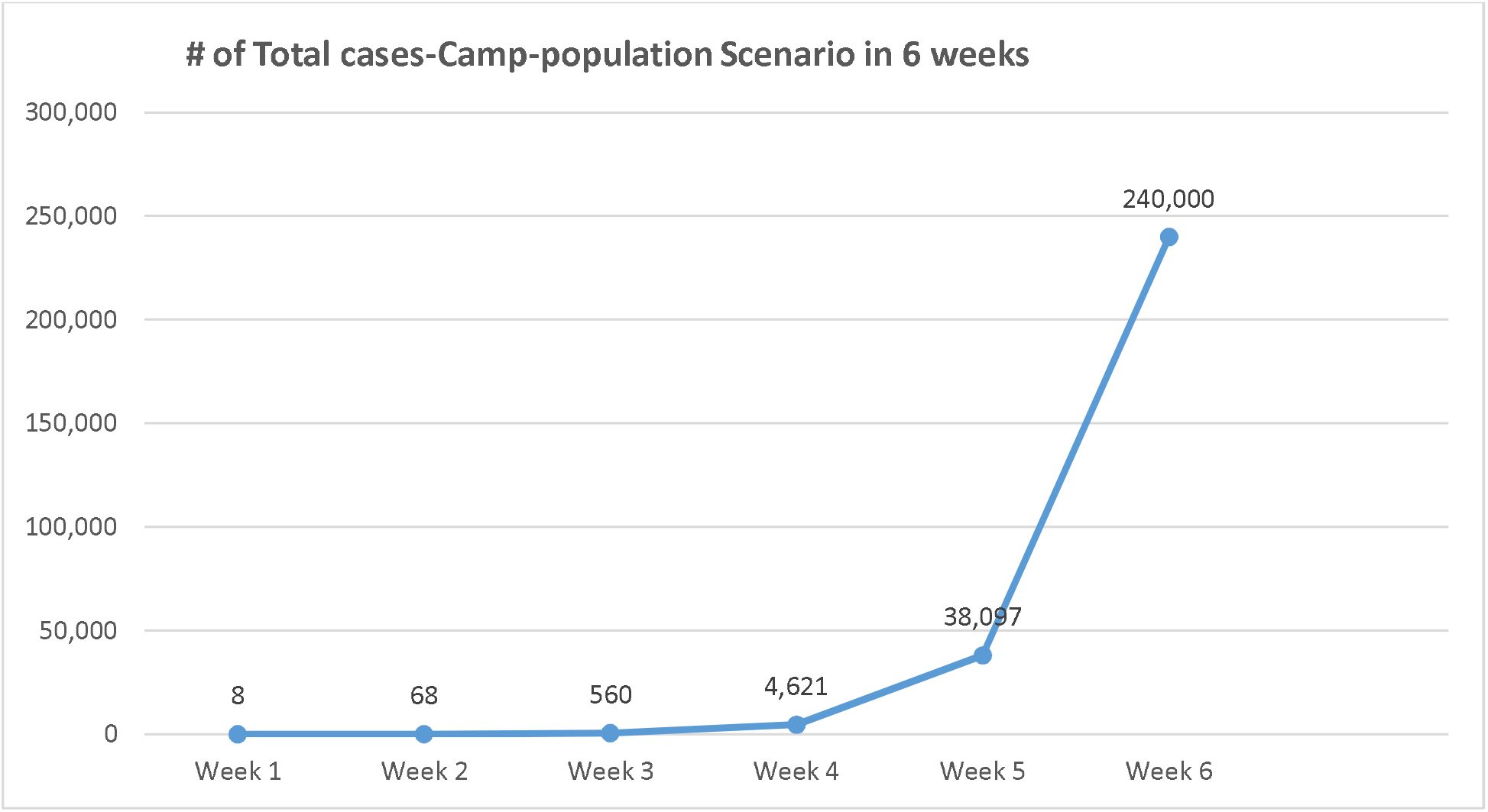
Camp-Population Scenario Predicted total cases

**Figure 15.**
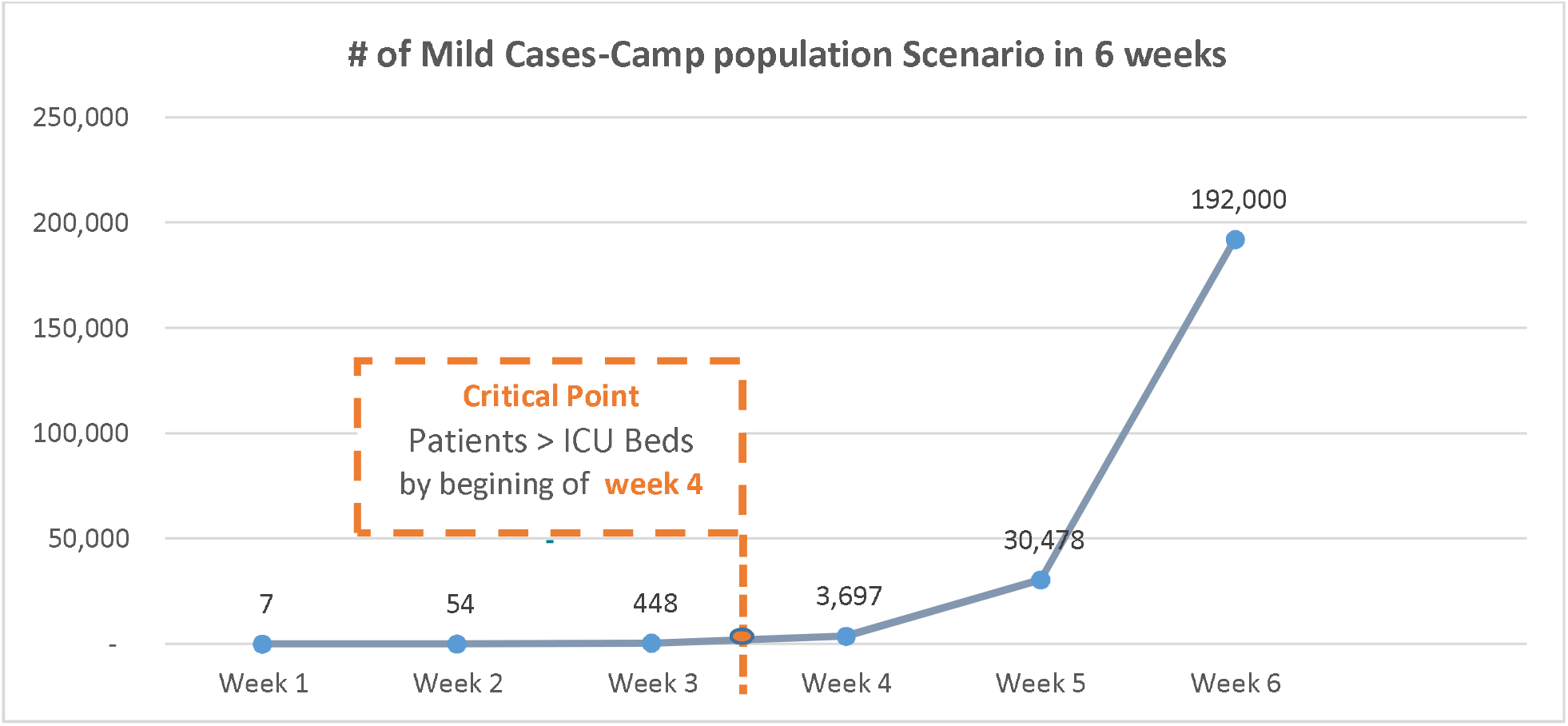
Camp-population Scenario predicted mild cases

**Figure 16.**
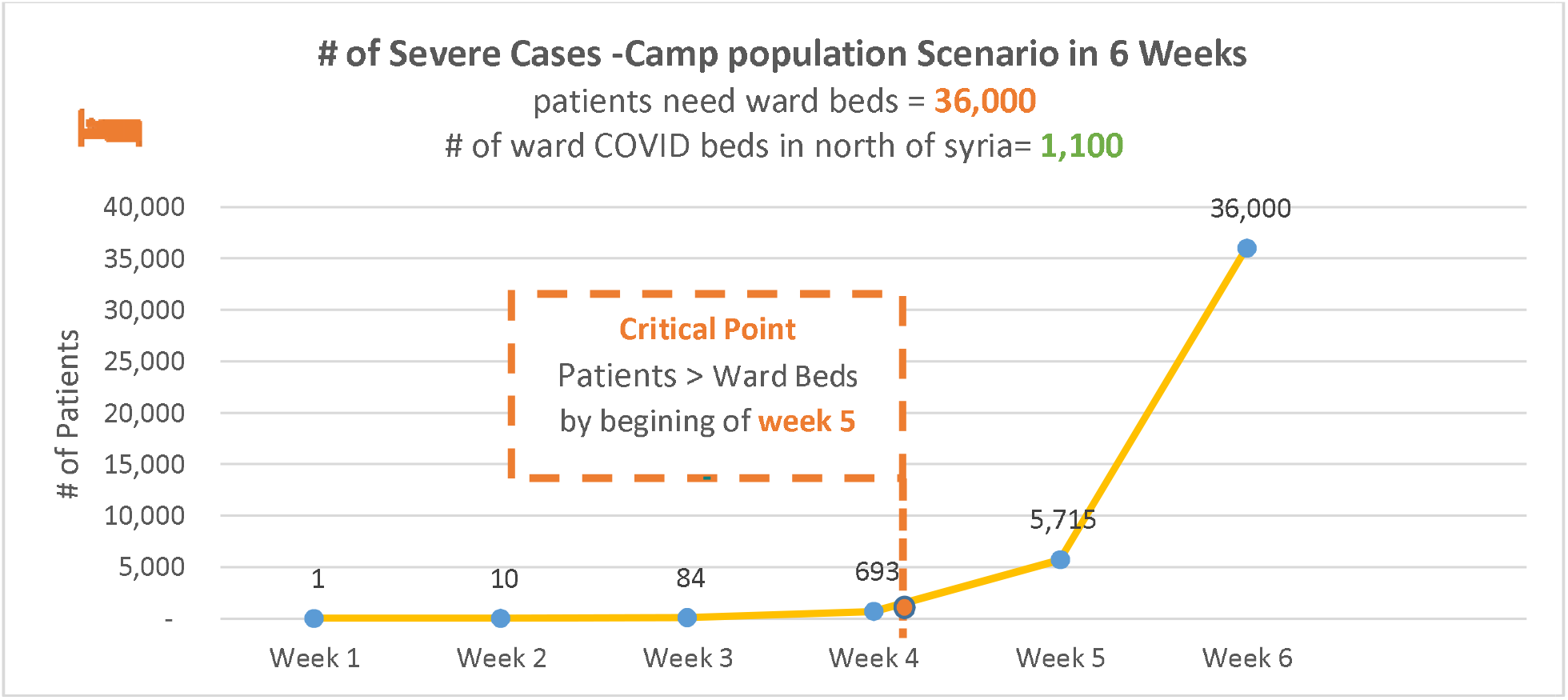
Camp-population Scenario predicted severe cases

**Figure 17.**
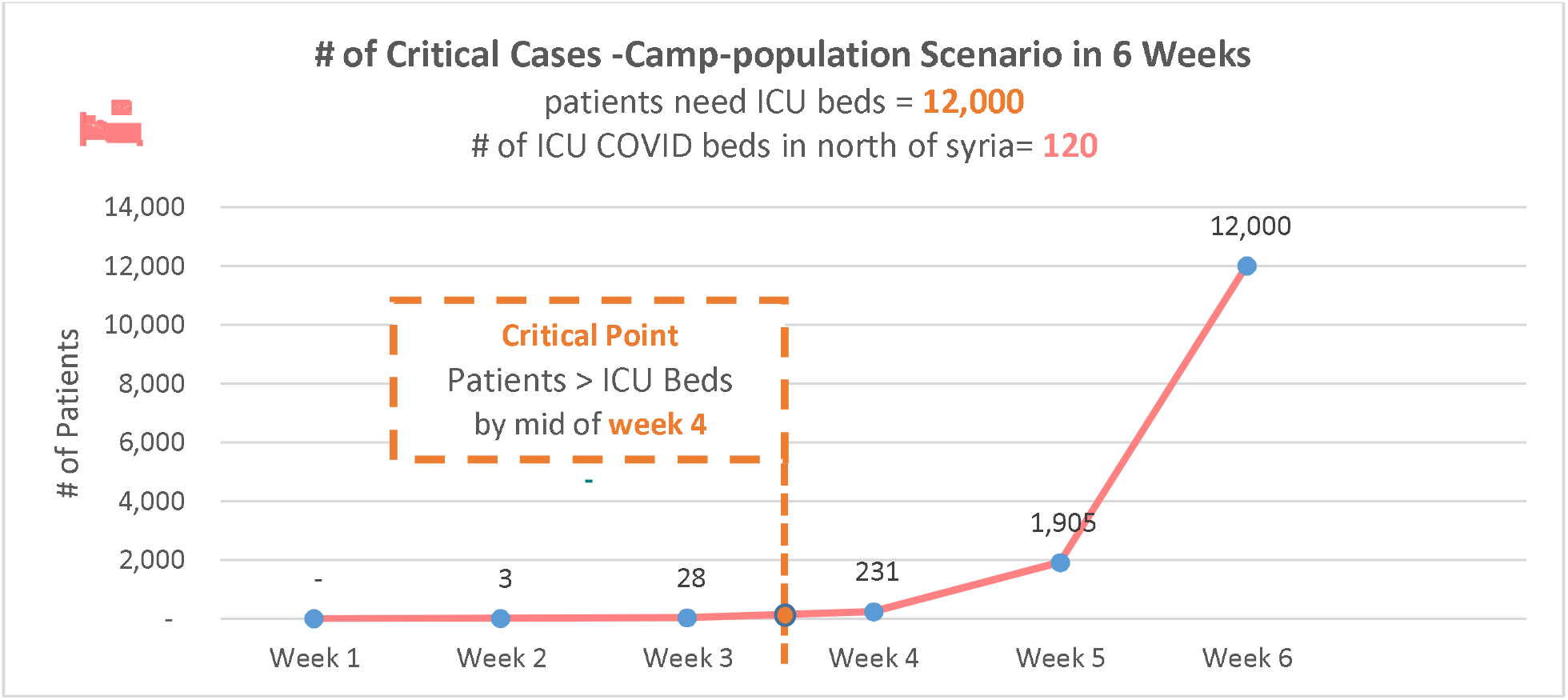
Camp-population Scenario predicted critical cases

**Figure 18.**
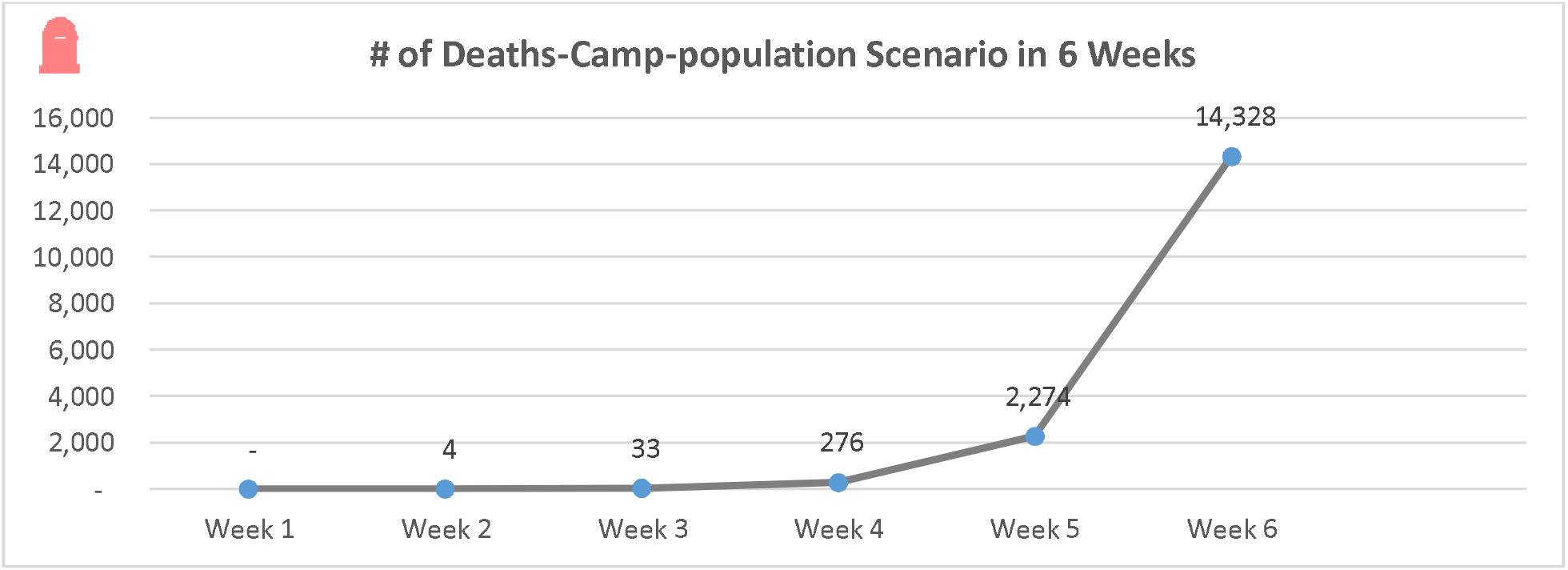
Scenario three predicted deaths

<u>Mild cases:</u> By the end of the 6 weeks forecasting period, 192,000 mild cases are expected which need to be identified and isolated. These cases could be also managed in CBI but CBI beds will be saturated very early on due to the sheer number of cases and the two weeks isolation period needed per patient.

<u>Severe cases:</u> The COVID-ESFT predicts that 35,000 out of total 240,000 cases will be severe. These cases need admission in inpatient departments of secondary health care centers but not ICU admission or ventilation. Needs for ward beds surpass available ward beds at the beginning of week 5, overwhelming the health system and leading to increases in the number of critical cases which could contribute to a higher mortality rate. Based on the current health system capacity**, <u>there is no possibility</u>** that the health system can manage this situation leading to its collapse.

<u>Critical cases:</u> Over 6 weeks, 12,000 cases will be critical, requiring ICU admission. The critical moment for critical cases in this scenario will be by the mid of 4^th^ week when ICU COVID-19 needs exceed 50% of available ICU beds capacity. <u>***It is impossible***</u> for the health system torespond to the expected numbers with its current capacity and the existing high occupancy rates (98%) of ICU beds.

<u>Deaths:</u> The COVID-ESFT predicts 14,328 deaths by the end of week 4. However, both the CFR and the number of deaths are likely to be much higher due to lack of the capacity of the health system to deal with the expected load of severe and critical cases.

To sum the figures about case load and expected critical time-points for health system capacity across the three scenarios, Figures 19 and 20 provide estimates for severe and critical cases, respectively.

**Figure 19.**
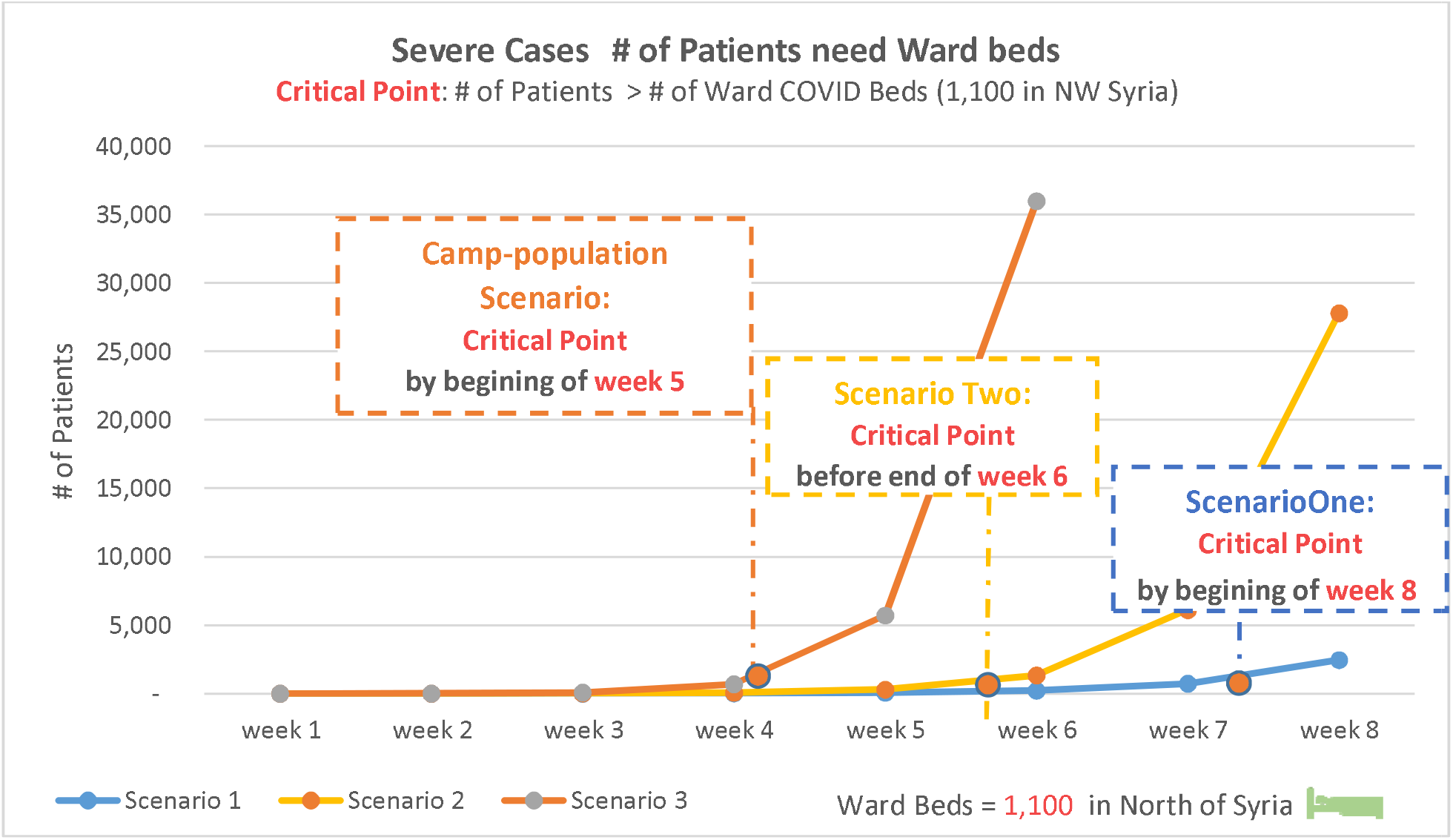
Comparison # of severe cases in all scenarios with critical points

**Figure 20.**
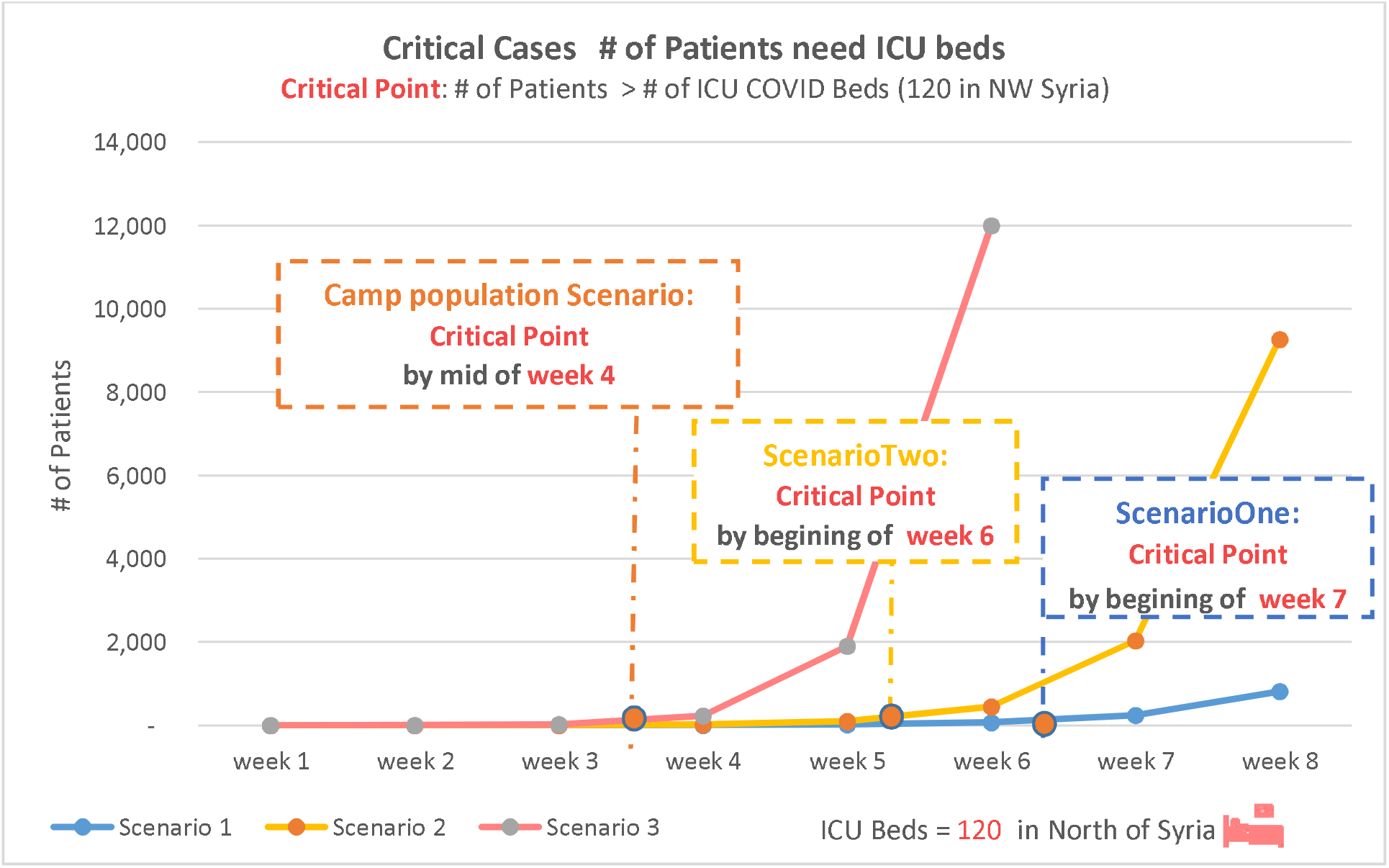
Comparison # of critical cases in all scenarios with critical points

## Discussion

Using a WHO forecasting tool, this study has identified COVID-19 case load, according to severity, and potential health system needs in NW Syria, an area that has been subjected to severe levels of violence, displacement and health system disruption during the nine years of the Syrian conflict. The study models use three scenarios. Scenarios One and Two apply to the total population of 4.17 million and for 8 weeks from the first case while the Camp-population Scenario applies only to the 1.2 million internally displaced persons (IDPs) in camps and tented settlements. For each scenario, we identify critical time-points when the health system capacity is overwhelmed assuming a highly conservative estimate that 50% of regular hospital (ward) and ICU beds can be occupied by COVID-19 patients.

*Scenario One* is better compared with Scenario Two but study researchers consider it unlikely for NW Syria given the density of the population, chronic exposure to severe stressors, and impracticality of implementing well-establishing measures of physical distancing and isolation. Even in this scenario, the health system would be unable to cope beyond week 7. *Scenario Two* is more realistic but is still not the worst-case scenario in NW Syria given the nature of overcrowding, inadequate shelter and poor access to WASH services. In this scenario, the health system is unable to cope beyond the fifth week. The Camp-population Scenario describes a situation whereby unmitigated spread in crowded displacement camps will lead to total health system collapse within the first four weeks of an outbreak. Considering that such a scenario can occur concurrently with Scenario Two (or a worse All-population Scenario Three not presented here whereby doubling rates and clinical attack rates would be higher), the epidemic outcomes can be catastrophic.

In all three scenarios, the projected time points of when the health system capacity gets overwhelmed assumes, conservatively, that 50% of the existing hospital and ICU beds and ventilators in NW Syria can be devoted exclusively to COVID-19. In reality, this would be extremely difficult considering high rates of occupancy of regular and ICU hospital beds (88%, 98%, respectively) and ongoing high levels of violence and injuries in NW Syria, meaning health system capacity is likely to be overwhelmed much sooner than the projected critical time-points. As COVID-19 patients require prolonged ventilator support,^22^ ventilators could become rapidly saturated with patients who are unable to be rapidly weaned. Our study did not account for this factor. It will be simply impossible to manage the predicted critical cases expected during the first eight weeks, in all scenarios, resulting in significant excess mortality.

We believe our projections hold true even acknowledging that non-COVID-19 presentations will fall during a COVID-19 outbreak, as he happened in other settings. Other countries took significant steps to decrease the bed occupancy rate by limiting admission for non-urgent or elective surgeries which allowed them to redirect more beds for COVID-19 patients. However, the baseline capacity of the health system in NW Syria is already significantly constrained. Therefore, there is very little extra capacity that can be gained by postponing non-urgent cases.

Mitigation measures being implemented in high- and middle-income countries in non-conflict affected settings are not feasible in NW Syria where insufficient WASH and shelter limit the implementation and effectiveness of social distancing, self-isolation, quarantine and lockdowns. There are also other risks of spread in crowded urban spaces especially during the holy month of Ramadan and Eid celebrations, lack of safety/protection should hostilities resume, lack of awareness and or interest in taking preventative measures by local population (i.e. given urgent situation outside of the pandemic). This could result in far greater and more rapid spread of COVID-19 among the population whose conditions are already poor. With insufficient numbers of community-based isolation, inpatient ward and ICU beds and ventilators, a COVID-19 epidemic in NW Syria could result in thousands of excess avoidable deaths. As a result of these factors, epidemic outcomes will be devastating. An additional concern compounding high numbers of deaths will be how to dispose safely of the bodies of those who have died due to COVID-19.

This study has several limitations that need to be addressed in future research. This forecasting report uses globally reported estimates for key parameters such as doubling rate, attack rates and case-fatality rates. It is possible for the situation in NW Syria to be different, either negatively or positively, thus substantially altering projections. This necessitates repeating these projections based on actual figures during an outbreak as has been done in other settings. Our study did not use age-specific projection (such as CFR). The COVID-ESFT does not incorporate population-specific factors, such as population age structure, prevalence of comorbidities particularly chronic conditions, and population density, and health system factors, such as bed availability and health workforce readiness, both of which can affect epidemic outcomes. As such, our findings should be considered basic and further modeling is needed to take such factors into account. Unfortunately, there is limited prior research on COVID-19 in similar conflict-affected settings^23^ to guide such modeling and epidemic projections. The COVID-ESFT considers an exponential growth model, which does not reflect a real epidemic curve that is seen in epidemics and pandemics. Furthermore, the COVID-ESFT only projects an estimate for the first six to eight weeks of an outbreak from the first case. Should an outbreak occur, real-time epidemiological data would be incorporated into the model to provide better forecasting. The study does not take into account the impact of important factors that can affect epidemic spread and outcomes: current preparedness and expected future interventions by the local authorities to control the spread of COVID-19 during the expected epidemic and the impact of community-based mitigation measures such as social distancing, self-isolation or shielding. More broadly, our knowledge about the SARS-CoV-2 virus is evolving; more understanding of its epidemiologic characteristics may alter this (and other) forecasting models.

## Recommendations

Based on the findings of this study, it is clear that there is a need for urgent action to mobilize efforts and resources to avoid a potentially catastrophic COVID-19 in NW Syria. Specifically, there is a need for innovative measures to limit human-to-human transmission, detect and isolate cases early and build sufficient capacity and resilience in the health system to mitigate excess mortality. NW Syria presents many challenges to such measures included an already overwhelmed and underfunded health system, insufficient testing, challenges of implementing internationally used approaches e.g. self-isolation, social distancing, strict lockdowns and a population affected by a protracted conflict of whom 67% are IDPs and around one quarter live in camps or tented settlements. Nonetheless, there are measures which can support the health system to prepare for the potential number of cases as forecasted in our modelling. We identify five priorities.

1. **Increase emergency funding** to support the health and humanitarian response in NW Syria.
2. **Upscale testing measures** including through support laboratory capacity including equipment, consumables, training and staff.
3. **Strengthen feasible community interventions**. With aforementioned limitations to implementing internationally-recommended guidelines, for example physical distancing, isolation, there is a need to support innovative approaches to limit community transmission. Several examples are illustrative. Implementing interventions targeted to local communities to increase physical distancing when possible; high-risk shielding of individuals, such as the over 60,000 people older than 60 years old and those with pre-existing morbidities; expanding camp capacity and preparing dedicated separate tent/s for isolation in each camp for isolation of suspected cases; improving access to water, sanitation and hygiene (WASH) measures in ways that are context-sensitive; ensuring that the public wear masks that are freely available but reserve N95 masks for aerosol generating procedures.
4. **Support and protect healthcare workers** through supporting training (infection prevention and control, skills for different healthcare facilities); availing PPE in sufficient quantities; support skill substitution; and providing staff compensation.
5. **Strengthen healthcare system capacity:** Ensure sufficient funding and support through the Health Cluster to support the health directorates and humanitarian organizations to build capacity including community isolation beds, inpatients beds with oxygen, ICU and ventilator capacity, expand testing (laboratory capacity and supplies, staff)

## Data Availability

This study uses WHO ESFT projecting tool to predict estimate of COVID-19 cases
This tool is publicly available
Estimate by study researchers can be made available on request

## Acknowledgements

Research team would like to express their very great appreciation to Dr. Aula Abbara^1^, Dr. Samer Jabbour^2^ Dr. Natasha Howard^3^, Dr. Hannah Clapham^4^, Dr. Abdulkarim Ekzayez^5^, Dr. Yazan Douedari for their valuable and constructive suggestions during the planning and development of this research work. Their willingness to give their time so generously have been very much appreciated.

^1^Consultant in infection and research fellow, Imperial College London; Chair, Syria Public Health Network

^2^Associate Professor of Public Health Practice, American University of Beirut ’s Faculty of Health Sciences.

^3^London School of Hygiene and Tropical Medicine and National University of Singapore

^4^Researcher, National University of Singapore

^5^Research for Health and Conflict in the Middle East (R4HC), King’s College London

^6^Syria Research Group, co-hosted by London School of Hygiene and Tropical Medicine and National University of Singapore School of Public Health

## Footnotes

1 https://gisanddata.maps.arcgis.com/apps/opsdashboard/index.html#/bda7594740fd40299423467b48e9ecf6

2 Case-Fatality Rate and Characteristics of Patients Dying in Relation to COVID-19 in Italy, Graziano Onder, MD, PhD1; Giovanni Rezza, MD2; Silvio Brusaferro, MD3, JAMA

3 https://coronavirus.jhu.edu/map.html

4 https://reliefweb.int/sites/reliefweb.int/files/resources/2019_Syr_HNO_Full.pdf

5 https://reliefweb.int/report/syrian-arab-republic/recent-developments-northwest-syria-situation-report-no-10-12-march-2020

6 https://reliefweb.int/report/syrian-arab-republic/recent-developments-northwest-syria-situation-report-no-10-12-march-2020

7 Health information System unit

8 HeRAMS 2020 Report, Turkey Health Cluster for Northern Syria, JANUARY - MARCH 2020

9 http://syriamap.phr.org/#/en/findings

10 IASC, set Global Health Cluster suggested set of Core indicators

11 Health Information System Team, DHIS2

12 HeRAMS 2020 Report, Turkey Health Cluster for Northern Syria, JANUARY - MARCH 2020

13 https://spherestandards.org/wp-content/uploads/Sphere-Handbook-2018-EN.pdf

14 Surveillance System for Attacks on Health Care (SSA), Syrian Arab Republic, annual report 2019

15 Health Information System Team, DHIS2

16 https://www.nature.com/articles/s41591-020-0822-7?fbclid=IwAR1Fpse5xcV__d75oBpnzOo0AhDMLHJnz0fZxkxW8cZPJXMkXmKRlJ6YfM%81j%82%E6%82%E8%81B

17 https://ourworldindata.org/coronavirus?fbclid=IwAR0N-V37byOhETU6xOCYGVkHJPasWZQ_OGeLaOORls1AZPRhTaktWDo6flU

18 Swerdlow DL, Finelli L. Preparation for Possible Sustained Transmission of 2019 Novel Coronavirus: Lessons from Previous Epidemics. JAMA. 2020;323(12):1129–1130. doi:10.1001/jama.2020.1960

19 https://www.thelancet.com/journals/laninf/article/PIIS1473-3099(20)30243-7/fulltext

20 By 20 April, the CFR had increased, but researcher decided to use the rate of 5.9% as a conservative estimate, given that all three scenarios are concerning even using a more conservative estimate.

21 Young BE, Ong SWX, Kalimuddin S, et al. Epidemiologic features and clinical course of patients infected with SARS-CoV-2 in Singapore.JAMA. Published March 3, 2020

22 https://www.thelancet.com/journals/lancet/article/PIIS0140-6736(20)30566-3/fulltext

23 Johns Hopkins, COVID-19: Projecting the impact in Rohingya refugee camps and beyond Summary

